# Care Giver’s Experiences of Having a Child with Hydrocephalus: A Phenomenological Study at Ruharo Mission Hospital

**DOI:** 10.1101/2020.06.25.20139683

**Authors:** Racheal Kyarimpa, Dan Muramuzi, Talbert Muhwezi

**Author notes:** Corresponding author; Talbert Muhwezi- Racheal Kyarimpa- Dan Muramuzi.

## Abstract

**Background:** Hydrocephalus is a long-term neurologic condition, normally identified in early childhood, where there is excessive fluid in the ventricular system within the brain which results into enlargement of the head to an abnormal size. The most common cause of hydrocephalus in most patient population is infection (over 60%) typically meningitis. In Uganda, it is estimated that about 1000 to 2000 new cases of hydrocephalus occur every year with 60 percent of these are reportedly attributed to neonatal infections. The general objective was to explore care givers‘ experiences of living with a child having hydrocephalus.

**Methods:** The phenomenological descriptive study involved fifteen respondents who were purposively selected. The in-depth interviews were conducted with the aid of an interview guide and a tape recorder. Transcribed interviews were analyzed using the thematic approach.

**Results:** Care givers were experiencing financial problems, psychological problems, lack of social support and cultural constraints and stigma associated with hydrocephalus. The care givers experiences were full of life changes and coping strategies, and they used both problem and emotion focused coping strategies to deal with the challenges as compassionate and responsive care was illustrated by the participants.

**Conclusion:** The study discovered that having a child with hydrocephalus is challenging and frustrating in terms of financial, physical, social, and psychological experience. Therefore, these findings are essential for counselling care givers, families and communities affected by hydrocephalus. In this context, attention should be targeted to families and communities to reduce stigma and isolation faced by the affected.

## Background

Hydrocephalus is a long-term neurological condition, normally identified in early childhood, where there is excessive fluid in the ventricular system within the brain which results into enlargement of the head to an abnormal size (Smith et al., 2013). Hydrocephalus is not a single disease entity but rather associated with developmental disorders such as spina bifida or encephalocele or may be due to brain tumors, head injuries and hemorrhage (Benjamin, 2005). It is also defined as an active distension of the ventricular system due to mismatch between cerebrospinal fluid production and its absorption (Jouibari et al., 2011). It represents one of the most common pathologic and neurological conditions affecting children requiring neurosurgical treatment (Abhaya et al., 2008).

The most common cause of hydrocephalus in most patient population is infection (over 60%) typically meningitis (Romero et al., 2014). Eventually, the child’s head will grow so big that the neck will not be able to support it. The child may not be able to sit or walk. The child is likely to become blind, deaf and mentally impaired (Aldersey, 2012). The child becomes more dependant in all spheres of life and care givers must find all the time required to look after such a child day after day (Rofail et al., 2013). This can be challenging to the care givers psychologically, physical, socially and economically (Civilibal et al., 2014).

Hydrocephalus is commonly treated by a surgical procedure, in which a tube called a shunt is placed into the child’s body known as Ventriculoperitoneal shunt (VPs). The shunt channels the flow of excess fluid away from the brain or spinal cord into another part of the body, where the fluid can be absorbed and transported to the bloodstream (Romero et al., 2014). Without a drain, the fluid buildup can affect brain development and cause disability or death as a result of malfunctioning shunt which is always the case.

The prevalence of hydrocephalus is estimated to be 400,000 new cases every year worldwide. In Sub-Saharan Africa more than 45,000 cases are seen every year, and in East Africa as a region, it is estimated that 6,500 new cases occur each year. In Uganda, it is estimated to be a range of 1000 to 2000 new cases occurring every year roughly, with 60 percent of these are reportedly attributed to neonatal infections (Bannink et al., 2015).

One recent large prospective multi-institutional study found that 40% of children who had VPs required a shunt revision within 2 years of initial shunt placement due to malfunction (Ogunleye et al., 2018). It was also found that 50% of the children who leave the hospital healthy after a successful surgery end up with blocked shunts resulting into child getting seizures, fever, headache among complications, thus making looking after these children more difficult (Benjamin, 2005).

More so, due to limited knowledge of most parents and neglect by immediate family members, the care givers of such children perceive their children as curses, having no value in the family and therefore not to be taken to hospital (Olive & Olive, 2007). Having understood that these children are dependent on their parents at all times. These care givers are faced with many challenges. Therefore, the aim of this study is to explore the care givers’ experiences of living with a hydrocephalus child.

## Methods

### Study design

This study utilized a phenomenological design to enable care givers recount their experiences, in order to allow issues that held most personal importance to them unfold. In depth interviews were undertaken to gain deeper insights using a face to face approach where the care givers were encouraged to direct and shape the discussion in accordance with their own experiences, views and particular concerns rather than responding to a pre-determined agenda. Discussions were held in a setting where participants felt comfortable to express themselves freely. The free interaction between the care givers and the researcher generated more information about caring for a child having hydrocephalus. The study was conducted between November 2018 and January 2019.

### Study setting

This study was conducted at OURS rehabilitation center, Ruharo Mission Hospital in Mbarara district. Ruharo Mission Hospital is a Private-Not for Profit Faith based institution of church of Uganda under Ankole Diocese. The hospital is located at Ruharo, 2 kilometers from Mbarara town on the Mbarara-Bushenyi-Fortportal highway, in Kamukuzi division of Mbarara Municipality. The OURS department happens to be the only rehabilitation center for children with hydrocephalus, Spina bifida, clubfoot, cleft lip/palate or deformities and other neurological disorders in the whole of western region. It also has both in-patient and out-patient departments and it rehabilitates over 500 children per year.

### Study participants

All the 15 study participants were purposively selected. Participants were considered eligible if they were having children with hydrocephalus and willingly consented to participate in the study. The care givers were excluded for this study if they were not able to finish the entire interview process.

### Data collection

Each eligible participant was required to first complete a consent form and a short demographic questionnaire before the interview. Individual in depth face to face interviews were conducted in a quiet private room. A research interview guide, note taker and tape recorder were used in this study. An interview guide with open-ended questions which were translated into Runyankore from English and back translated to its original form to ensure that the original meaning is not altered. This guide helped the interviewer pursue the same basic lines of inquiry with each person interviewed and managed the interviews in a more systematic and comprehensive way and to generate detailed and in-depth descriptions of human experiences. The researcher used a tape recorder during the interviews and after did the transcribing into verbatim because recording is more accurate than writing out notes. The interviews lasted between 45 and 90 minutes for each interview.

### Data analysis

Each interview was transcribed verbatim, and transcripts were analyzed using thematic analysis, facilitating exploration of how care givers ascribe meaning to their experiences while caring for their children. The analysis process began at the interview stage, with the researcher recording the thoughts feelings and responses to the interview process and content. Transcription further promoted familiarization with the data and generation of initial emerging themes. The transcripts were also analyzed in conjunction with the original recording, so that the researcher gets fully immersed in the data. Each transcript was coded and categorized, and an initial narrative summary of the key themes for in depth discussions made.

## Results

### Experiences of caring for a child with hydrocephalus

In the analysis of the lived experiences among care givers of children with hydrocephalus, four themes emerged. The four themes were: financial problems, psychological problems, lack of social support and cultural constraints and stigma. The four themes were interrelated and at times overlapping from the way the care givers described their experiences. Generally, the care givers experiences were full of life changes and strategies to cope.

### Financial problems

Most of the respondents experienced financial constraints while caring for children with hydrocephalus. This is due to the nature of the condition of the children they are caring for whose care requires one to be able to meet all the needs. These can range from basic needs, purchasing medical equipment to support the child, transport to and from the hospital, paying hospital bills as well as putting up structures that facilitate their movement. All these require a person caring for such a child to either have a high income or an investment to constantly rely upon to meets these demands.

*Money is the major problem in taking care of this child. I don’t have a job to give me income. I only use the little money I have to rent land for farming in order to raise more money to pay the hospital costs and buy some items to use at home. As a father, I feel exhausted and even, at times I have to get a loan in order to go to the hospital but the interest rates are so high. I was told that the surgery of my child requires 5 million shillings and for sure even if I sold everything in my house, I cannot get that money. They operated my son and they told me to pay shs. 950000 Ugandan shillings and they were to contribute for me the remaining money, but even the shs. 950,000 I never had it by that time so after the surgery I had to go back home and sell my land and up to now am still trying to clear the debt*. **(Father 2)**

Most respondents experienced the inability to meet other family basic needs like clothing, soap thus this caused them to feel irresponsible care givers.

*My husband got a fractured leg following an accident and he cannot move. At home, I am the one who struggles to look for money to meet the both family needs and hospital costs. I have many brothers who are rich, but they do not want know my situation. They cannot even buy me one bar of soap. For example, I currently owe the hospital a balance of shs1,700,000 for the surgical operation which was done earlier on my child. I just need God’s mercy because I see am still far*. **(Mother 5)**

*Haaa…*., *sometimes I don’t even have money to buy soap to wash the clothes for my child. My husband comes home only to sleep. He can’t buy soap, or even clothes for our daughter. My daughter cannot even accept to put on a torn cloth. So, every time she is asking for a new one, yet there is no money*. **(Mother 2)**

*At first, I really suffered because my husband was taking too much alcohol and every time I would go to Mbale (CURE Hospital), other children would be suffering because he would not give them scholastic materials to use, instead he sells everything and takes the money to the bar to drink with his friends*. **(Mother 6)**

*The main challenge is money because you find you are earning less or no income, yet his management requires a lot of money. Since his birth, I have struggled to raise the money to the extent of selling my land to meet the hospital costs. Sometimes you find that you have no transport but you sell like a goat so that you bring the child to the hospital. Also, at times you find that you don’t have money to buy drug supplements and maybe catheters because they are expensive. For example, whenever we are coming to the hospital, I have to buy a packet of pampers*. **(Father 1)**

*Uuhm, its alarming! transport to come here is also my challenge yet it’s always my wish to come for appointment, but still even the materials like catheters, medication to use while at home are always done by that time. So, I have to try by all means yet the father can’t send even the money for transport*. **(Mother 1)**

### Psychological problems

Most respondents experienced psychological problems in form of self-blame, negative perception from community and rejection by the spouse. Most mothers had experienced negative perception and rejection by their spouses and who married other wives after recognizing that they had delivered such children who required too much money and care hence leaving their wives alone to suffer.

*Hmm… when my husband left me alone in the village, I felt hurt and lonely because I had nothing to do*. **(Mother 1)**

*I had taken the child to the hospital on coming back I found when my husband had left with all his clothes as she breaks down in tears*. **(Mother 2)**

*The father of this child died and the mother had to take full responsibility of looking after this him. She got stressed to the extent that she sold everything including iron sheets on the house. Then one early morning at 6:00am she brought him on my door way and disappeared till now*. **(Grandmother 2)**

Some respondents experienced a feeling of self-blame because of the situation their children condition had positioned them.

*As my first born I felt bad and I always ask myself why me??***(Mother 2)**

*We reached at an extent of asking ourselves if we had committed any crime*. **(Father 2)**

However, some respondents reported improvement in self-esteem from the time they have been coming to the hospital because they have been counseled. Interaction with other parents with same problem has also transformed their lives.

*When I was home, I thought I was the only un lucky woman to produce such child but when I came to the hospital, I found we were very many and even others in worse situations*. **(Mother 3)**

*When we reached Mbale and saw many parents with children with the same condition I felt I was not the only one even when we reached home and told them, they changed positively their perception*. **(Father 2)**

Most respondents experienced negative perception from the community because the attitude of people surrounding them was not good. Some of them were treated as outcasts, and they would not associate with the rest of the community.

*Wherever I would pass with my child, people would stare at me and laugh. I would really feel like an outcast in the community*. **(Father 1)**

*After delivery, his mother dumped him here and she left because she could not bear taking care of such a child. Some children would not come to visit us because they never wanted to associate with my son he used to move as he is defecating on himself*. **(Grandmother 1)**

*Eeeh…, what people say makes one lose the morale of looking after the child including people at home*. **(Mother 5)**

*When I looked at this child. I could not understand what happened and what to do to him. I thought of throwing him away. But when you look at him, you just say let me try to do what I can as a mother*. (**Mother 1**)

However, one respondent hoped that with time the community‘s perception would change since these days they no longer segregate them as it was in the past.

*After sometime, some of my village members started to understand the agony I was enduring and they would give help in terms of money, food and clothes*. **(Guardian 2)**

Most respondents experienced fear and anxiety about the outcome of surgery, complications of the disease and mistreatment of their children.

*When they told me that they were going to operate my child, I got scared. I was are not sure whether my child would still be alive after surgery. I was seated next to another mother and she told me that three things may happen as he comes from theatre, he may be having fever, cough or dead. I got scared and I was really worried*. **(Mother 7)**

*After the operation, the doctor called my name. I remember my husband and I thought our daughter was dead. I started sweating and my husband refused to go there but I gathered all the courage as a mother and went there*. **(Mother 4)**

*…. Sometimes, people would tell me to leave my child to die. I would get angry but keep quiet and cry from inside. I would comfort myself that my child will die when I have done my part of taking her to the hospital*. **(Mother 6)**

Most respondents experienced fear that their children at school would be mistreated by their fellow pupils because there was no one to monitor them for their safety. And this made other parents hesitant to take these children to school.

*Am always anxious that any time that other children at school will beat him up or knock his head down*. **(Guardian 1)**

*My son has not gone to school because I fear that he will be mocked by the normal children at the school because of his abnormal shape of the head*. **(Father 2)**

*When he is with other children, they call him all abusive words like you deformed person, don’t you know yourself, and can’t you see how your head looks like*. **(Grandmother 3)**

Some care givers experienced fear about the complications especially having recurrent convulsions whereby it affects the normal functioning of these children. This puts them at tension whether it’s a shunt malfunction or an infection which has set in.

*Every time I see my girl convulse, I get so anxious and feel as if she is going to die. I get scared that one of the convulsions will be severe and she won’t recover*. **(Mother 4)**

*One day I asked the nurse whether my child would remain with the tube in his head or it will be removed. This is because it makes me worried especially when he gets sick, I start asking myself if its blocked? Or there is another problem*. **(Grandmother 3)**

### Lack of social support

Care givers mainly experienced lack of support either from their spouses, relatives and family members as a result of the child’s condition. This impacted their social aspects in terms of emotional, instrumental, appraisal and informational aspects.

*My wife no longer helps me in the garden because she can’t leave this child alone*. **(Father 1)**.

*My husband used to sell off everything I left at home and would go and take alcohol. My children would suffer and fail not go to school. Even as I planned to go to the hospital, I would struggle to look for the money alone. Because my husband and the relatives would ask me why I am taking the child who is already dead to the hospital but I insisted. (****mother 6***)

*When my husband discovered that our child had this problem, he looked for money to take us to the hospital and we left him at home. After coming back, he decided to pack his things and left me with the child. But after sharing with some of my community members what happened to the child, they tried to mobilize some support for me. Two weeks ago, someone told that me that he is married to another wife and they stay in the nearby trading centre. It has been two years since he left and he has not sent us any support. (****Mother 1***)

*My wife was operated while delivering this child and so they stopped her from doing any heavy job so she does most of activities at home in addition to look after this child*. **(Father 2)**

*I have many brothers who are rich, they have money but they don’t want to know my problem. People of the world will love you only when you have no problems. Also, my husband got a fracture of the leg. He is also at home seated and is also another problem. I am the woman and the man of the home. Every time I have to carry this child on my back whenever I doing house chores because he can’t seat, and as a result my chest is always paining*. **(Mother 5)**

Respondents experienced hindrance to development of the family because every little money earned cannot be invested on a large scale because you are not sure of what is going to happen in the next hour.

*Every time am moving to hospital for different follow up, sometimes my son gets admitted and can’t do the work am supposed to do*. **(Father 2)**

However, some participants reported that their families were always supportive in all situations for the betterment of their children regardless of the situation.

*For us at home, we plan for this girl like any other child. We love and cherish her. In fact, she is dad’s girl. Whatever she requests from dad, he provides because we don’t want her to get stressed. My husband has really been my leaning shoulder in taking care for this child. We work together to make sure we plan and raise the money for transport and hospital costs for the next appointment date*. **(Mother 4)**

*…*.*my husband is very supportive. Whatever I request for he provides so that our child can be well. (****Mother 7***)

### Cultural constraints and stigma associated with hydrocephalus

The care givers expressed mixed reactions on what could have caused the hydrocephalus to their children. The condition was tied to different cultural traditions with some care givers attaching it to the curse or witchcraft done by the parent. Some of the care givers had never heard about the disease condition that their children had developed and therefore associated it to superstition.

*At the time of delivery, I was in the hospital with my husband. He was the one taking care of me. When the midwife explained to us the baby’s condition, we were able to understand what happened. But the family members had not even heard of hydrocephalus and they thought it was as a result of witchcraft or an abomination. My in laws reacted to me harshly because they had never seen it and thought I had brought witchcraft in their family. They labelled me a witch*. **(Mother 4)**

*For sure, I had never heard about it. But when I told people in our village about the condition of my child, they thought it was a curse in return to the sins we unknowingly committed. The people in my village could not call him a child but rather say that we committed a crime. Even now, whenever I take him to the nearby health centers, some people see him and run away*. (**Father 1)**

*…. at first, I thought my child was bewitched and I took my child to different traditional herbalists. I can tell you they took a lot of my money but the head kept on increasing in size. Later on, someone told me to try modern medicine and that’s how I got here*. **(Mother 5)**

*We had never heard of such a condition. After delivering this child, we thought it was a curse. We went to a health center and they told us that they have never seen such. Whenever the fellow villagers would come to check on us, they would say this is an animal. When he started to crawl, he had fecal incontinence and he would leave feaces wherever he would pass. All the children in our village would say we cannot go to that home because there is a child that defecates anyhow. Others would say, really when they look at that child, do they see any hope in him? (****Grandmother 1***)

*…. people in my village would tell me not to waste my time taking my child to the hospital. But when I took my child to the hospital, some also people looked at her told me that my daughter is not suffering from a medical condition but rather she was bewitched. I tried to explain to the Doctor and he told me not to listen to their stories. I also tried using herbal remedies from a traditional herbalist who gave me confidence that the remedy was highly effective for such conditions. He gave the remedy for smearing around the body and those for chewing and told me that in one week my child will be okay, but I did not tell the health workers*. **(Mother 6)**

## Discussion

The findings of our study show that care givers of children with hydrocephalus are full of challenges and coping strategies. This study has generated important information on the way in which care givers’ experience care. Whereas this study was undertaken in one hospital, these findings are likely to be of interest to all providers of in-patient care, as the themes and issues highlighted here may also resonate with those care services. The four main themes that will steer the discussion are: financial problems, psychological problems and lack of social support and cultural constraints and stigma.

Our study found out that the care givers faced a challenge of financial costs incurred either in paying hospital bills, transport to the hospital or buying food and other items during their stay in the hospital. Our findings concur with a study conducted in Brazil which revealed that mothers having children with hydrocephalus experienced financial difficulties (Ben et al., 2013). In addition, Bannik et al., (2015) found out that care givers of children with disabilities continue to experience financial problems. Similarly, Bourke- Taylor et al., (2014) found that children with complex care needs necessitate direct costs on families that include extra medical attention, equipment, technology, devices, medications, and specialized therapy services, as well additional costs such as costs related to modifications of the family home. Furthermore, existing literature shows that having a child with a disability interferes with the care givers ability to get employed, thus exerting a lot of pressure on finances (Ambikile & Outwater, 2012). The findings of our study are in line with a study conducted by Diana et al., (2013) found that care givers also experience additional financial burden including reduced income. Nonetheless, care givers in our study reported that they did whatever they could to ensure their children were well cared for and have a good life. Whenever possible, care givers relied on their own finances to pay for certain services and support for their children. Previous research has also revealed that in order for these children to participate in the opportunities available in daily life, care givers will use their own finances to make up for shortfalls related to technology, equipment, and resources (Bourke- Taylor et al., 2014).

Our findings indicate that having a child with hydrocephalus can have a detrimental psychological effect like low esteem to move with such children in communities as expressed by most parents. Care givers had a number of psychological effects ranging from self-blame, negative perception and rejection by the spouse, negative perception by the community which can lower their self-esteem and make them feel like their children were outcast. The findings of our study agree with a study done in United Kingdom which found that parents of children with hydrocephalus experienced significant psychological stress (Joanna et al., 2013). Similarly, Oti- Boadi, 2017 revealed that parents having children with disabilities reported experiencing psychological stress. More so, the psychological experiences mentioned by care givers in this study resonate with other studies, which showed that they are always burdened by the psychological challenges (Cramm & Nieboer, 2011; Norlin & Broberg, 2013). Similarly, other studies found out that care givers complained about feelings of sorrow and grief which originated from their dealings with other people’s frequent messages of negativity, hopelessness, anticipation of the child dying and being treated by others as if they were an outcast in the family (Olive and Bwana, 2007; Ayse et al., 2015).

Throughout this study, care givers expressed feelings of fear and anxiety due to multiple sources like; not knowing what will happen to their children, whether they going to live a normal life or not as they grow up and become adults? Other stressors were whether their children would live with shunts in their heads for life, if they would require revisions as their children grew or whether the shunt would be permanent throughout adulthood. In another study, the intense parenting also resulted in health challenges for parents, including parents experiencing physical problems and increased stress, anxiety, and depression (Woodgate et al., 2015). Similarly, Joan et al., (2013) found out that worry and anxiety were part of daily life and dominated care givers’ accounts of living with a child with hydrocephalus.

This study revealed that care givers experienced deteriorating social support from their partners, friends and society. Other research studies have reported that mothers of children with hydrocephalus experience enormous burden as a result of the limited support they receive from their family and the community (Aldersey, 2012; Gupta et al., 2012). Mothers need to be supported emotionally and financially by families, friends, society, health professional, governmental, and nongovernmental agencies concerned with improving the lives of mothers and their children with hydrocephalus. The necessity to have great support from family and society to successfully raise their children was a concern for care givers in this study. Mothers with strong support from their spouses, siblings, grandparents, and their children’s school reported improved psychological health. Existing research emphasizes the important role of support from family, friends, church, and professionals in alleviating stress and facilitating positive coping ability among parents of children with hydrocephalus (Aldersey, 2012; Ha et al., 2011; McNally & Mannan, 2013). However, most of the care givers have found their hope in Christ for their children’s future and this has helped them cope up with stress and anxiety. Moreover, care givers in this study reported that they received support from their faith-based leadership and congregation. This resonates with existing literature which indicates that support provided by religious organizations improves the psychological health of parents of children with hydrocephalus (Pillay et al., 2012).

Our findings show that care givers experienced cultural constraints and stigma associated with having children with hydrocephalus, since they were perceived as outcasts, witches or curse in their families and society. This could be as a result of little knowledge and superstitious beliefs held by the family members, relatives, friends and society in the African tradition. Our results compare with what has been reported from another study in Uganda by Bannink et al., (2016) which found that due to little knowledge of most society members and mistreatment of parents of such children, they tend to take these children as curse, or a product of witch craft. Therefore, they find these children having no value in the family, hence not to be taken to the hospital and even parents are always stigmatized. The spiritual interpretation of disability in the African context may have also influenced care givers’ interpretation of their children’s condition. Oti-Boadi, (2017) asserted that Black mothers of children with congenital problems could not tell the cause of their children’s condition due to their external locus of control of attributing their children’s situation to supernatural causes. Clearly, the findings of this study showed that mothers’ experiences are situated in the context of how they believe society views and treat their children (Baffoe, 2013; Hervie, 2013). However, research has shown that in Africa, health beliefs have been described as holistic, where many families and communities hold multiple beliefs, consisting of medical and African traditional and supernatural belief system explanations and treatment of diseases (White, 2015).

## Conclusions

The findings from this study show that care givers of children having hydrocephalus live a challenging life. These care givers live a life of financial, psychological, social and cultural challenges. Compared to other qualitative studies on care givers experiences of having a child with hydrocephalus, our results give a detailed account of the psychological and social challenges care givers face. Therefore, it is important to have ongoing psycho-social counselling to encourage care givers not to lose hope on providing care to their children.

### Study Limitations

This study was not without limitations. First, findings from this study cannot be generalized to all care givers of children with hydrocephalus in Uganda. The present study focused on care givers living in one region of the country; therefore, it remains unknown whether care givers living in other regions differ in their lived experiences. Future studies into the experiences of health workers may provide a rich understanding into the dynamics of caring for a child with hydrocephalus. Nevertheless, findings from this study provide a suitable guide for future research into the experiences of parents and may lead to an improvement in their holistic well-being.

### Implications for Interventions and Policies

Emphasis should be placed on empowering families with coping strategies associated with their cultural values to help them traverse the challenges associated with raising children with hydrocephalus. Mental health professionals should support care givers in considering the positive aspects of their experiences, offer them hope, assist them to make efforts in reframing their situation, and make meaning of the experience of having a child with hydrocephalus. Findings of this study could inform the design of culturally sensitive programs and policy on children with hydrocephalus. Financial support from government and other corporate organizations for mothers to educate their children and support themselves should be provided.

## Data Availability

Data available from the corresponding author at a reasonable request

## List of abbreviations

CSF: Cerebrospinal fluid
ETV: Endoscopic third ventriculostomy
FGD: Focused group discussion
HCWs: Health Care Workers
LMIC: Low Middle-Income Countries
OURS: Organized useful rehabilitation services
RMH: Ruharo mission hospital
UHP: Ugandan Hydrocephalus Project
VPs: Ventriculoperitoneal shunt
WHO: World Health Organization

## Ethical approval and consent to participate

Ethical approval was obtained from the Bishop Stuart University- Research Ethics Committee. All participants provided informed written consent for participation in the study after an explanation about its objectives, confidentiality, and ethical consideration, and assurance concerning the voluntary nature of participation.

## Consent for publication

Not applicable.

## Availability of data and materials

The data used to elicit the findings of this study are available from the corresponding authors upon reasonable request

## Competing interests

The authors declare that they have no competing interests

## Funding

Not applicable

## Author’s contributions

RK, TM, DM participated in the conception and design of the study. RK and DM conducted the interviews and TM participated in data analysis. TM wrote the manuscript and RK and DM contributed to the drafting of the manuscript. All authors read and approved the final manuscript.

## Acknowledgements

The authors would like to express their sincere thanks to the care givers who agreed to share their experiences in the interviews and FDGs. Thanks to Ruharo Mission Hospital for allowing us to interact us with their patients through their care givers. We would also like to appreciate Ms. Rachel Luwaga for her constructive guidance during concept development.

## Notes

### Competing Interest Statement

The authors have declared no competing interest.

### Funding Statement

No external funding was received

### Author Declarations

BISHOP STUART UNIVERSITY RESEARCH ETHICS COMMITTEE

